# Forecasting Cycles of Seizure Likelihood

**DOI:** 10.1101/2019.12.19.19015453

**Authors:** Philippa J. Karoly, Mark J. Cook, Matias Maturana, Ewan S. Nurse, Daniel Payne, Ben Brinkmann, David B. Grayden, Sonya B. Dumanis, Mark P. Richardson, Greg Worrell, Andreas Schulze-Bonhage, Levin Kuhlmann, Dean R. Freestone

## Abstract

**Objective:** Seizure unpredictability is rated as one of the most challenging aspects of living with epilepsy. Seizure likelihood can be influenced by a range of environmental and physiological factors that are difficult to measure and quantify. However, some generalizable patterns have been demonstrated in seizure onset. A majority of people with epilepsy exhibit circadian rhythms in their seizure times and many also show slower, multiday patterns. Seizure cycles can be measured using a range of recording modalities, including self-reported electronic seizure diaries. This study aimed to develop personalized forecasts from a mobile seizure diary app.

**Methods:** Forecasts based on circadian and multiday seizure cycles were tested pseudo-prospectively using data from 33 app users (mean of 103 seizures per subject). Individual’s strongest cycles were estimated from their reported seizure times and used to derive the likelihood of future seizures. The forecasting approach was validated using self-reported events and electrographic seizures from the Neurovista dataset, an existing database of long-term electroencephalography that has been widely used to develop forecasting algorithms.

**Results:** The validation dataset showed that forecasts of seizure likelihood based on self-reported cycles were predictive of electrographic seizures. Forecasts using only mobile app diaries allowed users to spend an average of 62.8% of their time in a low-risk state, with 16.6% of their time in a high-risk warning state. On average, 64.5% of seizures occurred during high-risk states and less than 10% of seizures occurred in low-risk states.

**Significance:** Seizure diary apps can provide personalized forecasts of seizure likelihood that are accurate and clinically relevant for electrographic seizures. These results have immediate potential for translation to a prospective seizure forecasting trial using a mobile diary app. It is our hope that seizure forecasting apps will one day give people with epilepsy greater confidence in managing their daily activities.

## INTRODUCTION

Medically refractory epilepsy is a condition associated with persistent uncertainty. Most people with epilepsy report that, regardless of seizure frequency, it is the unpredictability of when seizures will occur that is the most debilitating aspect of their condition ^1^. A forecast of seizure likelihood could provide immense benefits to people with epilepsy, especially for those with refractory seizures. Over 30% of all people with epilepsy cannot control their seizures with medication ^2^ and, for decades, new drugs have mostly failed to improve overall rates of seizure freedom ^3^. Surveys have shown that seizure forecasting is considered a highly attractive management option by people with uncontrolled seizures ^1^. At the same time, epilepsy research has uncovered numerous factors affecting seizure likelihood from long-term trends in behavioural, environmental and physiological data. For example, some people are more prone to seizures due to stress ^4^, poor sleep ^5^, exercise ^6^, diet ^7^, weather ^8^, alcohol use ^9^, poor drug adherence ^10^ and a multitude of other factors. Investigating the utility of these seizure risk factors and developing personalised forecasting devices is now a key goal for clinical epilepsy management ^1^.

The first prospective clinical trial of seizure forecasting in humans used an intracranial implant to record long-term, continuous electroencephalography (EEG) ^11^. Using the same data, subsequent studies have improved the accuracy of seizure forecasts ^12,13^. A key development has been the understanding that seizure onset is modulated by patient-specific, cyclic patterns ^14–16^. The rhythmic nature of epilepsy has been well documented for centuries; however, it has only recently become clear that an individual’s seizure cycles can be used to develop personalized forecasts of future seizure likelihood ^13,14,17,18^. Although changes in EEG signal characteristics provide the clearest biomarker to track cycles of seizure likelihood ^16,18,19^, it is possible to measure individual cycles using only self-reported seizure diaries ^15^. Therefore, for some people, seizure diaries alone may provide a clinically useful forecast of future high or low seizure risk periods.

It is now recognised that estimating the probability of someone having a seizure in the near future is more feasible than trying to predict the exact timing of their next seizure ^12,17,20^. There are increasing efforts to develop a clinical seizure forecasting device ^1^ and understand user requirements ^21–23^. Patient surveys have confirmed that probabilistic forecasts are considered useful and that perfect accuracy is not a requirement of such devices ^21,23^. For practical reasons, externally worn devices are rated more desirable than implanted recording devices in surveys ^23^. There is increasing availability of wearable technology for seizure detection and forecasting ^24–26^. Furthermore, many seizure triggers that have been shown to be useful biomarkers of seizure likelihood can be measured non-invasively. For instance, self-reported stress is predictive of seizures ^27,28^. Heart rate ^29,30^ and other physiological signals monitored from a smartwatch device ^31^ have also been used to forecast seizure likelihood. It is possible that these non-cerebral biomarkers of seizure likelihood are useful because the same fundamental rhythms that modulate many aspects of human physiology also drive seizure risk.

This study used long-term, self-reported data from a mobile seizure diary to determine whether seizure cycles can provide a useful forecast of future seizure likelihood. The results provide a proof-of-concept that forecasting seizure cycles is practical and accurate and has the potential to be used as a clinical management tool. Mobile tools to forecast seizure cycles have wide-ranging applications including for improved clinical trial design, treatment titration and long-term management for people with epilepsy.

## METHODS

### Data

This study used long-term data from a mobile seizure diary app (Seer Medical) to develop and test seizure forecasts. Forecasts were also generated using only the self-reported events from the Neurovista dataset, an existing set of long-term, continuous EEG and seizure annotations that has been widely used to develop forecasting algorithms ^11^. The Neurovista dataset enabled forecasts based on self-reported seizures to be evaluated against electrographic seizures, and also provided a comparison of non-invasive forecasting performance to state-of-the-art seizure prediction.

#### Mobile app data

The Seer app is a freely available mobile diary for reporting seizures and medications. Currently, there are over 500 active users. This study used a subset of 33 users with a clinical epilepsy diagnosis, at least 30 reported seizures (mean 103 seizures), and at least 2-months recording duration (mean 42 weeks). Although users can report information such as seizure type and duration, this study only used the times of reported seizures. This study was approved by the St Vincent’s Hospital Human Research Ethics Committee (HREC LRR 165/19).

#### Continuous EEG data

The Neurovista study collected continuous intracranial EEG data from 15 subjects for a period of 6 months to 2 years. Further information on data collection and participants are reported by Cook et al (2013). This study only used the seizure times reported during the 6 month – 2 year recording periods. Electrographic seizures were automatically annotated by an onboard EEG-based detection algorithm. All seizure detections were confirmed by trained epileptologists. Self-reported events were based on diaries kept by participants and caregivers, which were subsequently combined with EEG annotations and stored as electronic records. In addition to diary data, the Neurovista device included an audio recording feature that was automatically activated when suspect epileptiform EEG activity was detected by the onboard algorithm. The audio recordings were used to aid confirmation of clinical seizures; and, for some subjects, audio recordings provided substantial assistance in seizure detection. However, to provide a direct comparison with mobile app data, the current study primarily considered events based on participants’ diaries. The human research ethics committees of the participating institutes approved the Neurovista study and subsequent amendments. All patients gave written informed consent before participation.

### Forecasting seizure cycles

An overview of the forecasting method is shown in Fig. 1. An individual’s strongest circadian (“fast”) and multiday (“slow”) cycles were estimated from their reported seizure times using the magnitude of the resultant vector, or synchronisation index (SI), measured over a range of potential cycle periods. The synchronisation index is given by:

**Figure 1.**
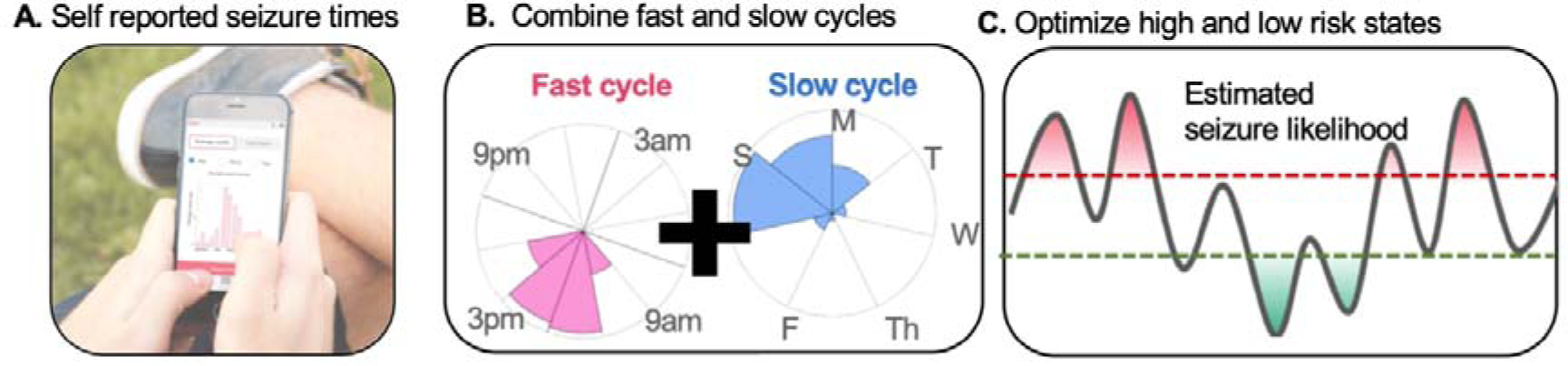
Schematic of seizure forecasting method. **A**. Development of forecasts initially required at least 20 seizure times to be reported in the mobile app. **B**. Self-reported seizure times were represented as cyclic histograms with different periods. The strongest fast and slow cycles were estimated based on the synchronisation index of seizure histograms. In the example, a 24 hour and 7 day cycle are shown. **C**. The estimated seizure likelihood was calculated by combining the fast and slow cycles. High and low risk forecasts (dashed red and green lines) were optimised based on historical data.

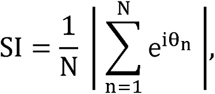

where N is the total number of seizures and each seizure is represented as a vector on the unit circle, 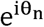. The angle, θ_n_, ranges from 0 to the period of the cycle being estimated; i.e., 24 hours for a circadian cycle. For this study, fast cycle periods were assumed to be between 6 hours and 48 hours, with a 6-hour increment; i.e. the strongest cycles were selected from 8 candidate periods (6,12,18, 24, 36, 42 and 48 hours). This range of fast cycle periods enabled the detection of approximate daily cycles rather than true circadian rhythms. Slow cycles were assumed to have periods of greater than 3 days, with a maximum allowed period of 2 months or 1/5 of the recording duration (whichever was lowest). For instance, a person with 10 weeks of data could have a maximum slow cycle of 2 weeks, whereas someone with 1 year of data could have a slow cycle of up to 2 months. The increment for slow cycles was 1 day; i.e. the strongest cycles were selected from candidate periods of [3 days, 4 days, 5 days, …] up to the maximum allowed period. Only significant cycles were used to develop seizure forecasts. Significance was assessed using the Rayleigh test for non-uniformity of circular distributions (p < 0.05) ^32^.

Individuals’ strongest cycles were iteratively updated with each new seizure based on an exponentially weighted history of past seizure times; i.e., the sum for the synchronisation index of N seizures became:

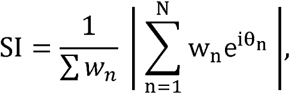

where w_n_ = 1 − 1 − α ^*n*^ is the weight applied to the n^th^ seizure and α = 0.8. To estimate seizure likelihood, the phases of the fast and slow cycles, ϕ_slow_ and ϕ_fast_, were derived for each hour over the recording duration:

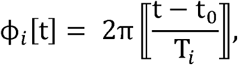

where T is the period of the strongest cycle (i.e. T_fast_ or T_slow_ and ⟦·⟧ indicates the modulo operator (the remainder after division). The phase, “is a function of time, t (hours), and t_0_ represents the time of the most recent seizure (as the estimated fast and slow cycle periods, T, were updated after each seizure). The phase was then quantized into 20 equally spaced bins, from 0 to 2π. The probability of seizure occurrence with respect to each phase, P [ϕ_slow_] and P [ϕ_fast_], was calculated from the histogram of previous seizure times.

The final probability of seizure occurrence at each time, P[t], was obtained as the product of the log-odds of each probability, P[ϕ_slow_] and P[ϕ_fast_]^33^.

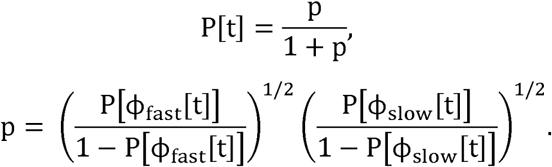

High and low risk warning thresholds were computed using a pseudoprospective brute force optimization that maximized the time spent in low risk periods and number of seizures classified in high risk periods ^18^.

Forecasts were updated every hour and evaluated pseudoprospectively, using only the historical seizure record to compute the likelihood of future seizure occurrence. At least 10 seizures were required to initiate the seizure forecast, which was then iteratively updated after each seizure. Only the lead seizure within each 1 hour window was used, subsequent seizures within the same hour were removed before calculating and evaluating forecasts. Performance was evaluated based on the percentage of seizures in high (low) risk and the total duration spent in high (low) risk, after setting optimal thresholds for high and low risk warnings. A 5 minute intervention period was used for evaluation; i.e., seizures were only considered to be correctly predicted if the high risk warning was on for at least 5 minutes before onset. Note that because seizure likelihood was calculated hourly, this intervention period effectively meant that if a seizure occurred less than 5 minutes past the hour, the high risk threshold needed to be exceeded in the previous hour to be considered a true positive. Seizures more than five minutes past the hour were assessed as true positives if the high risk threshold was exceeded within the same hour. Sensitivity improvement over chance was calculated based on the proportion of seizures and time in high risk to assess whether the high risk forecast sensitivity was significantly better than chance performance ^34^.

Performance was also measured using the receiver-operating characteristic (area under the curve, AUC). The AUC addresses the ability of a classifier to discriminate between inter-ictal and pre-ictal data and is the preferred measure for many studies benchmarking multiple seizure forecasting algorithms ^34^. The AUC was also used to assess the significance of forecasting performance compared to chance level forecasting. Significance was tested using 95% confidence intervals of the AUC using 1000 bootstrap replicas (using *perfcurve* in MATLAB). The AUC was compared between actual and random forecasts using non-overlapping confidence intervals (a conservative measure) and the one-tailed t-test (p < 0.05).

## RESULTS

### Can people self-report their seizure cycles?

We had previously demonstrated that seizure cycles can be measured from self-reported seizure times ^15^. The current study presents a retrospective validation that self-reported cycles correspond to true underlying epileptic rhythms, rather than just forecasting behavioural cycles governing when individuals are more or less likely to report their seizures. The Neurovista dataset was used to determine whether self-reported seizure cycles aligned with cycles derived from electrographic seizures.

Fig. 2 shows that 11 out of 15 Neurovista subjects (73%) had circadian cycles that were not significantly different using self-reported (diary) events compared to electrographic seizures (p > 0.05 using Kuiper’s test for circular distributions). Three subjects (S2, S3, and S5) did not have enough diary events to make a comparison. Nine subjects (60%) had multiday cycles that were not significantly different using diaries compared to electrographic seizures (see supplementary Fig. S1). This suggests that although participants vastly underreported the total number of seizures, the underlying circadian and multiday trends can still be determined. There was a preference for individuals to report less seizures at night (supplementary Fig. S2). In comparison, audio confirmations were higher at night and lower during the day, perhaps reflecting less background noise during the night. In general audio confirmations improved the alignment of circadian rhythms (supplementary Fig. S3). Overall, these data suggest that self-reported seizures identified clinically relevant cycles for most people with epilepsy, albeit based on a relatively small validation cohort with refractory, focal epilepsy.

**Figure 2.**
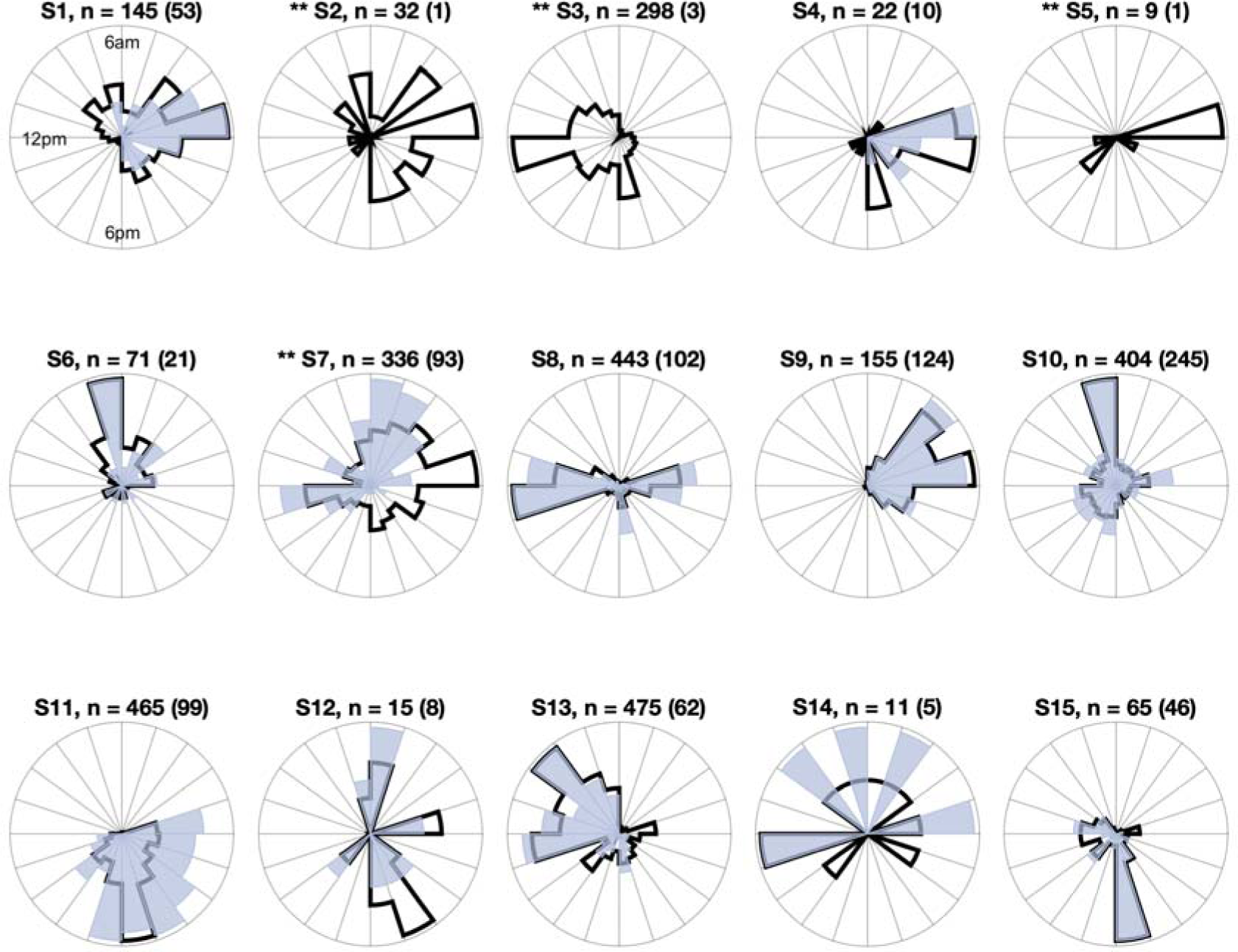
Alignment of seizure cycles based on self-report and EEG for the Neurovista cohort. Each circular histogram shows the circadian distribution of seizure cycles based on self-reported events (light blue) and electrographic seizures confirmed by expert reviewers (black line). Total seizure numbers for each subject are given above each plot (with self-reported event counts in brackets). Subject 7, had significantly different cycle distributions using Kuiper’s test for circular distributions (* p < 0.05, ** p < 0.01). Subjects 2, 3 and 5 did not have enough self-reported events for a comparison to be made.

Fig. 3 shows a comparison of forecasting performance using either self-reported (diary) or electrographic seizures from the Neurovista cohort. Both forecast models were evaluated using electrographic seizures. This comparison provided a unique opportunity to benchmark the performance of forecasts based on self-reported seizures against confirmed electrographic seizures, ensuring that forecasts were not merely useful at predicting users’ reporting habits. This benchmarking is critical to provide confidence in the clinical utility of forecasts based on mobile diaries alone.

**Figure 3.**
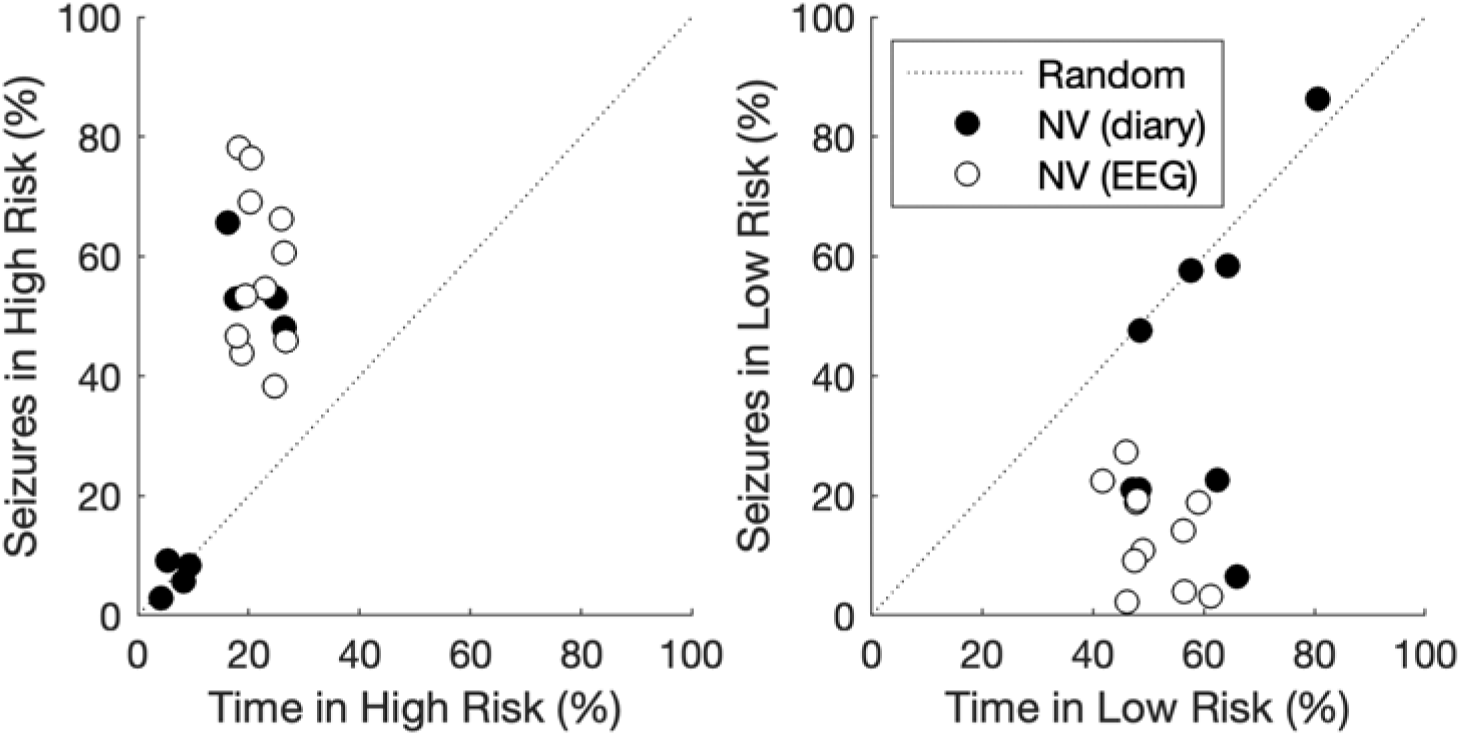
Validation of forecasting performance based on self-reported seizure cycles. A comparison of forecasting accuracy using self-reported events (black) and electrographic seizures (white) from the Neurovista cohort. Note that only individuals with at least 30 seizures were included. Forecasting performance was evaluated for electrographic seizures. **A**. Percentage of time in high risk (x-axis) compared to accuracy, or proportion of seizures reported during high risk states (y-axis). A good forecast is near the top left corner (100% accuracy, with the minimum number of hours in high risk). **B**. Percentage of time in low risk (x-axis) compared to the proportion of seizures in low risk states (y-axis) reported. A good forecast is near the lower right corner, indicating maximal time in low risk without any seizures occurring.

Forecasts were developed for subjects with at least 30 seizures (11 subjects had >30 electrographic seizures, eight subjects had >30 self-reported seizures). Using electrographic seizures to develop forecasts resulted in a mean time in high risk of 20.5%, with 59.5% of seizures occurring in this state. The mean time in low risk was 55.6%, with 13.0% of seizures. However, when just self-reported seizures were used to develop forecasts, several individuals showed a marked decline in forecasting performance. Based on self-reported forecasts, four of the eight Neurovista subjects showed performance not significantly different from random performance (p > 0.05 using the sensitivity improvement over chance metric) for high risk warning states. The other half of the subjects fell solidly within the low or high risk clusters. This result shows that forecasts based on self-reported events may provide an accurate forecast of the likelihood of clinical electrographic seizures for approximately half of the cohort, while the other half of this group would produce inaccurate self-report forecasts.

### Forecasting accuracy

We tested whether a seizure forecasting model developed from self-reported seizures could be used to forecast future self-reported events using the mobile app diary data. Fig. 4 shows an example of what forecasting seizure likelihood looked like using data recorded via the mobile app. The individual shown had an average of 2.8 seizures per week. Over the year of pseudo-prospective evaluation, their forecast showed 18.7% of the time in high risk, with most seizures (59.3%) reported in this state. The individual’s forecast showed 55.1% of the time in low risk and 10 of their seizures (8.9%) occurred during low-risk periods. The rest of the time (26.2%) was spent in the moderate risk state. Supplementary Fig. S4 shows two other examples of subjects with different forecasting outcomes. The supplementary examples show that diary forecasts can provide useful information for individuals with a range of seizure rates, including an individual with a lower reported rate of 1.4 seizures per week who had no seizures occurring during low risk periods, as well as a subject with a high rate of 24.2 seizures per week.

**Figure 4.**
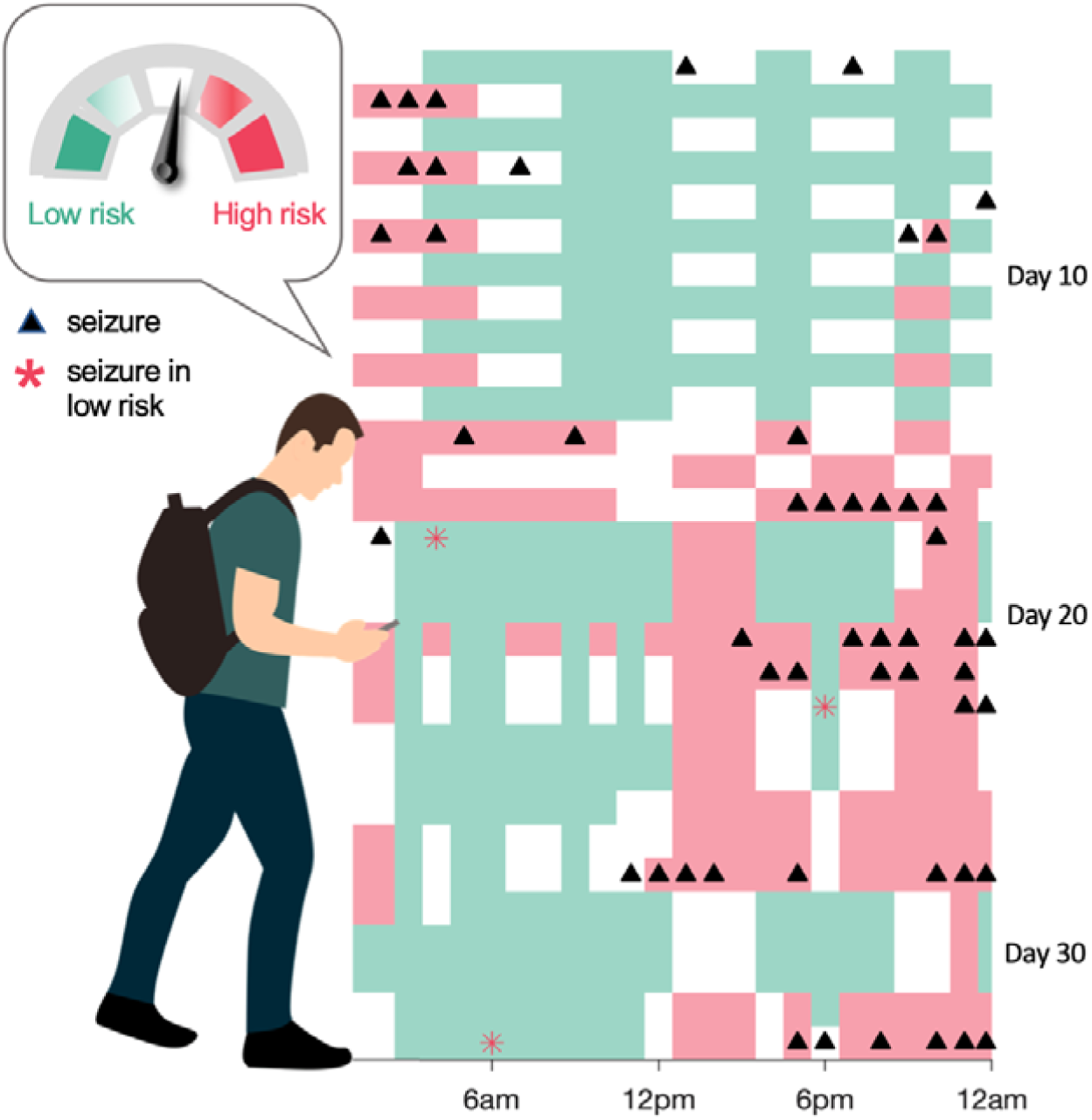
Output of a seizure forecast. Example data from an individual’s high and low risk forecasts over a 30-day period (y-axis). The panel shows the pseudoprospective forecast output each day (x-axis). Red indicates times when the high risk warning would be activated, green indicates time when the low-risk warning would be activated and white shows times when the warning was moderate. Reported seizures are marked in black, and seizures that occurred during the low risk state marked as red asterisks (in this data three seizures occurred during low risk periods). Note that individuals were not shown the output of their forecasts, and these data represent an example of pseudoprospective results only.

Fig. 5 shows forecasting performance based on mobile app data. It can be seen that using mobile diaries, the mean high risk accuracy was 64.5% with users spending a mean of 16.6% of their time in high risk. The mean time in low risk was 62.8% with an average of 9.7% of seizures occurring during low risk states. Seven of the 33 app users (21%) had zero reported seizures during their low risk periods. All forecasts were significantly better than chance (p < 0.05 using the sensitivity improvement over chance metric). It is important to note that these results were evaluated based on self-reported seizures and may not reflect the performance if all electrographic seizures were recorded. However, the results suggest that clinically useful forecasts can be developed from mobile data alone, although it is anticipated that, for approximately half of people, mobile forecasts will not be accurate for predicting electrographic seizures, as shown from the Neurovista validation cohort (Fig. 3).

**Figure 5.**
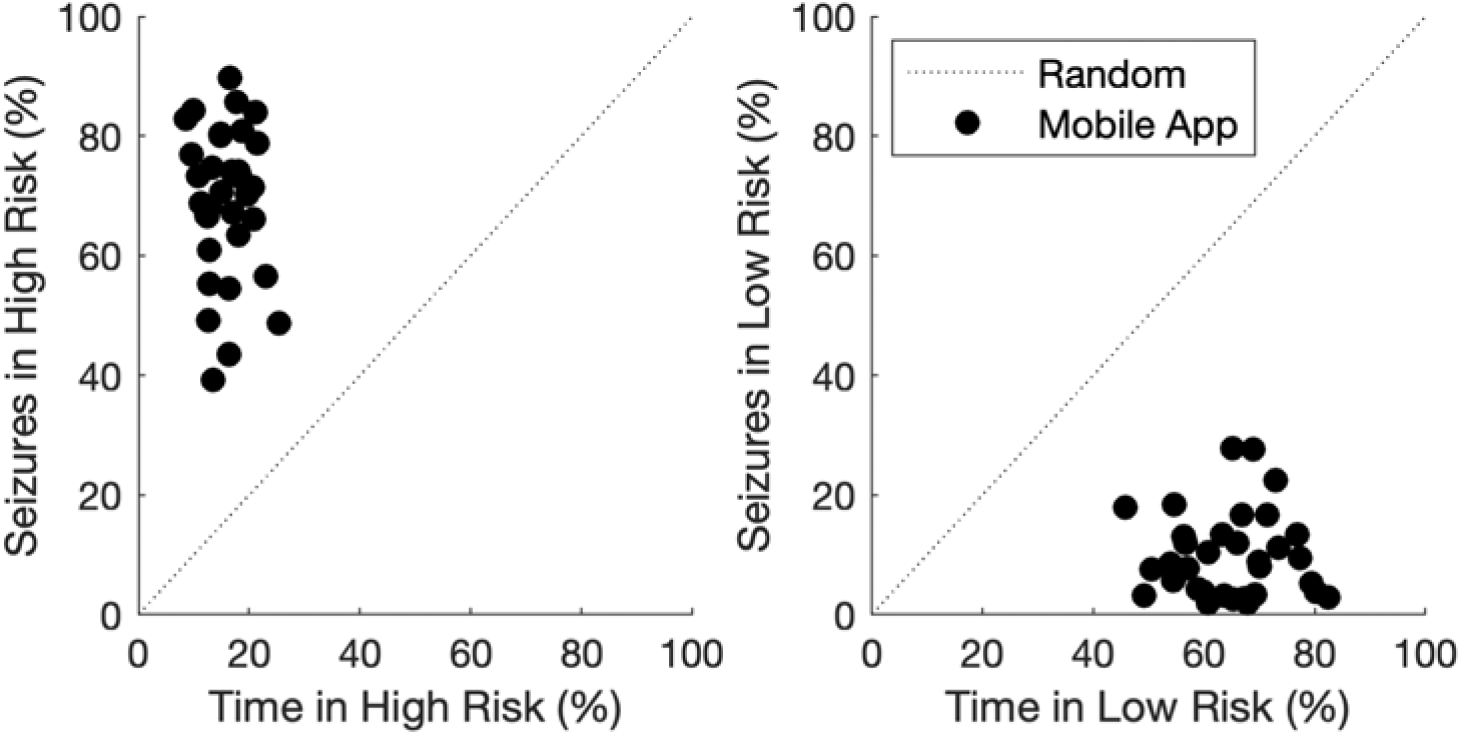
Forecasting performance using self-reported seizure cycles. Performance of high and low risk forecasts developed from mobile app data for 33 users. **A**. Percentage of time in high risk (x-axis) compared to accuracy, or proportion of seizures reported during high risk states (y-axis). A good forecast is near the top left corner (100% accuracy, with the minimum number of hours in high risk). **B**. Percentage of time in low risk (x-axis) compared to the proportion of seizures in low risk states (y-axis) reported. A good forecast is near the lower right corner, indicating maximal time in low risk without any seizures occurring.

Fig. 6 shows the rate of true positive compared to false positive predictions for different thresholds of high risk warnings using mobile app data. Area under the curve (AUC) provides a measure of forecasting performance without explicitly setting a warning threshold, where AUC greater than 0.5 indicates performance is better than chance level. Importantly, 95% confidence intervals can be computed for each AUC value, to show whether results were significantly better than a random forecast. Across the cohort, the mean AUC was 0.85 (range of 0.69 to 0.94). All individuals had AUCs that were significantly better than chance performance (95% confidence intervals were non overlapping and p < 0.05 using a two sample t-test).

**Figure 6.**
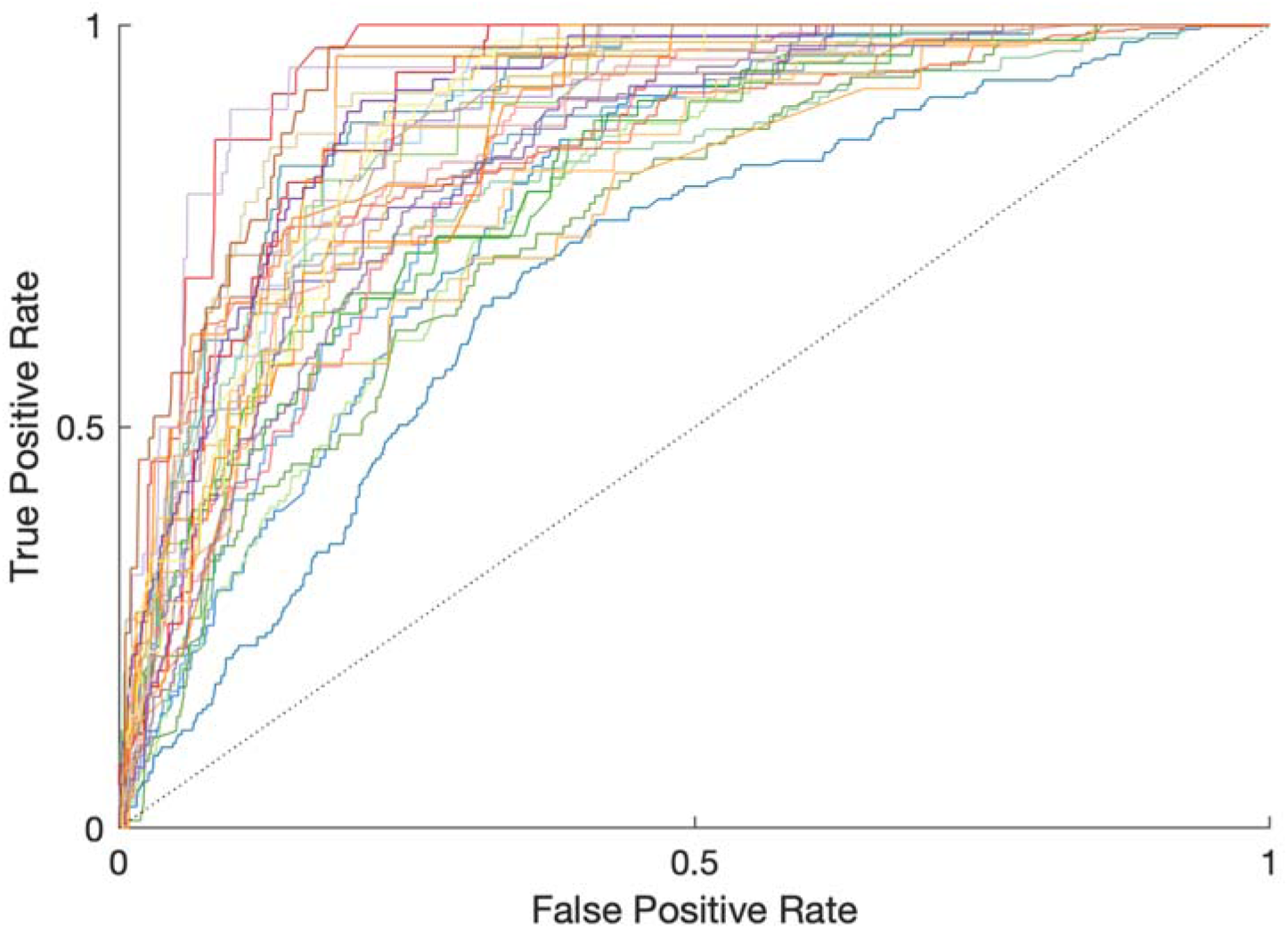
Receiver operating characteristic curves for seizure forecasts. The curves show the true positive rate (y-axis) compared to the false positive rate (x-axis) for each individual app user as the high risk threshold is varied. An ideal forecast has AUC = 1; chance level forecast has AUC = 0.5 (the diagonal line). The 95% confidence intervals of each curve were computed using a bootstrap analysis of 1000 permutations. Note that, for clarity, the confidence intervals are not plotted although these were used to calculate the significance of AUC scores.

## DISCUSSION

It is well known that seizure diaries are inaccurate and not well correlated to true seizure rates ^11,36^. The presented results showed that, although diaries are an unreliable estimate of true seizure counts, they can be used to measure underlying epileptic rhythms (Fig. 2). Because seizure cycles are repetitive, diary data can be treated as a noisy, undersampled representation of these underlying patterns. In this way, many people’s seizure rhythms were reliably measured from their diaries and could be used to forecast clinical electrographic seizures (Fig. 3). Underreporting and a slight bias towards daytime reporting did not abolish overall cyclic trends, and the use of audio confirmation increased the number of recorded seizures and improved the estimation of seizure cycles (supplementary Figs. S2 and S3). Some subjects reported almost no seizures, which could reflect many possible factors, including post-ictal memory impairment, the available social support, or the subject’s level of engagement and attention to detail during the study. On the whole, the retrospective validation of self-reported forecasting performance against electrographic seizures (Fig. 3) suggested that seizure forecasting apps will be accurate and may be a clinically useful method for some people. Future work will focus on incorporating objective biomarkers of seizures from wearable devices and validating the clinical utility of the proposed seizure forecasting app in a prospective clinical trial.

Overall the presented results showed that seizure diary apps have the potential to provide accurate, clinically useful, personalized forecasts of seizure likelihood. Forecasting results using seizure diaries showed state-of-the-art performance, with an average AUC of 0.85 (Fig. 6). A recent Kaggle competition for seizure prediction using continuous intracranial EEG recorded from three subjects in the Neurovista cohort reported winning AUC results of 0.81 for the competition and 0.75 on the held-out dataset ^13^. Of course, these scores are not directly comparable to app forecasts, as they were derived from continuous intracranial EEG data with highly accurate seizure labelling; however, the comparison serves to highlight what is considered top range performance in seizure prediction using advanced machine learning. The current study showed forecasting accuracy of 65%, with app users spending, on average, less than one fifth of their time with a high risk warning (16.6%). A study using deep learning with the Neurovista data reported an average sensitivity of 69% and time in warning of 27% ^37^. Moreover, forecasts based on seizure cycles naturally provide insight into times of low seizure likelihood. In this study, users were able to spend, on average, over half their time in a low risk state. Less than 10% of reported seizures occurred during the low risk state and 20% of users had no seizures in this state. Of course, it should be reiterated that this study only evaluated performance from users’ self-reported events. It is anticipated that around half of the cohort would only show chance level performance for their electrographic seizures (Fig. 3). However, the same forecasting strategy showed similar forecasting performance for both diary data and electrographic seizures (Figs. 3 and 5). Therefore, performance is expected to improve as more accurate records of individuals’ seizures becomes available, for instance through new wearable or implantable technologies. Furthermore, there are applications for forecasts of self-reported events, such as scheduling EEG monitoring, or improving analysis of clinical trial diaries ^38^.

Delivering seizure forecasts in a prospective trial is itself non-interventional in the sense that no direct therapy is given. Instead, users are only given information about their personalized seizure likelihood. However, reliable information may be a powerful antidote to the uncertainty of living with uncontrolled seizures. Furthermore, information about seizure likelihood can be used in conjunction with behavioural strategies to reduce seizure rates. For instance, it has recently been shown that stress management techniques could be targeted to days of heightened seizure risk to reduce seizure rates ^39^. Forecasts may also be used to modulate medication or stimulation levels based on seizure likelihood. Such modulating chronotherapy has been used to time dosage of anti-seizure medication, successfully reducing seizure rates ^40^. However, modulation of drug levels has not been investigated for individually tailored cycles or over longer timescales to account for multiday cycles of seizure likelihood. Seizure forecasting systems may also improve seizure detection and prediction accuracy by incorporating data from wearable or implantable monitoring devices.

Wearable devices have long been heralded as the next frontier in epilepsy management, both for their potential as automated seizure detectors and to provide advance warning of seizure onset ^24^. However, wearable seizure monitors have faced several challenges, including their relevance for more subtle seizure types, high false alarm rates and poor user experience ^22,41^. A forecasting app could potentially improve the detection performance of wearable devices by providing a prior probability of seizure likelihood. Similarly, physiological signals recorded from wearable devices could improve forecasts of seizure cycles by providing a continuous measure of underlying rhythms, rather than discrete samples (seizure times). We have shown that fast and slow cycles of brain activity can be measured from continuous EEG across diverse frequencies and regions of cortex ^18^. These continuous cycles provided the most accurate estimate of seizure likelihood to date. More recently, cycles of interictal epileptiform were also used to provide a forecast of seizure risk over days ^19^. It is possible that cycles of epileptic brain activity can also be derived from auxiliary systems modulated by the brain, such as cardiac and pulmonary output or even mood and sleep. It is well known that both circadian and multiday cycles modulate many aspects of human health and disease ^42–44^, including heart disease ^45,46^, immune response ^47^ and neurological and psychiatric disorders ^48,49^. However, a long-term study to identify cyclic rhythms of physiological signals in conjunction with cycles of seizure risk has not yet been undertaken. Ultimately, the ability to measure cycles from almost every biological process underscores the power of a simple, cyclic seizure forecast.

There is still no consensus on how accurate a seizure forecast must be in order to be clinically useful, nor for which epilepsy characteristics and patterns forecasting methods may prove most useful. Several studies have surveyed the views of people with epilepsy and caregivers on the subject of seizure forecasting. In one survey, participants reported that missed seizures were considered worse than false alarms, and that perfect accuracy was not considered a requirement for a forecasting device ^21^. Recently, Janse et al. (2019) showed that seizure forecasting devices were acceptable despite the potential for inaccuracy (up to “inaccurate 30% of the time”). Ultimately, post-hoc studies and surveys can only provide an indicative measure of the utility and benefits of a seizure forecasting device. In a prospective setting, some people may find knowing times of safety to be more important than high risk warnings. Some people are likely to tolerate false alarms poorly, whereas others would find a forecasting device very helpful. As with many aspects of epilepsy treatment, the usefulness of seizure forecasting is likely to be patient specific.

This study provided a proof-of-concept that seizure diary apps can provide a personalized forecasts of seizure likelihood. We hypothesise that mobile apps to forecast cycles of seizure likelihood have the power to improve quality of life for people with epilepsy, even without delivering direct intervention. In support of this hypothesis, we note that unpredictability is the primary disability of epilepsy ^1^ and, for people with refractory seizures, quality of life is far more strongly determined by the degree of drug-related side effects or depressive symptoms than by seizure frequency ^50^. Reliable seizure forecasting has the potential to improve quality of life by reducing uncertainty and improving mood. There is also scope for forecasting to reduce drug related side effects through intelligent titration of medication. Ultimately, it is our hope that seizure forecasting apps provide another clinical tool to manage epilepsy.

## Data Availability

Raw data were provided by Seer Medical (mobile app data) and Neurovista (EEG seizures). Derived data supporting the findings of this study are available via a data sharing agreement by request to the corresponding author. The Neurovista Kaggle competition and seizure data are accessible online via https://www.epilepsyecosystem.org.

## ACKNOWLEDGEMENTS

This project was funded by the National Health and Medical Research Council Project Grant (APP 1130468). This project was also supported by the Epilepsy Foundation of America’s Epilepsy Innovation Institute “My Seizure Gauge” grant.

MPR is funded in part by the MRC Centre for Neurodevelopmental Disorders, the EPSRC Centre for Predictive Modelling in Healthcare, and the NIHR Biomedical Research Centre at the South London and Maudsley NHS Foundation Trust.

ASB receives study-related funding from My Seizure Gauge and from the RADAR-CNS project.

## SUPPLEMENTARY MATERIAL

**Figure S1.**
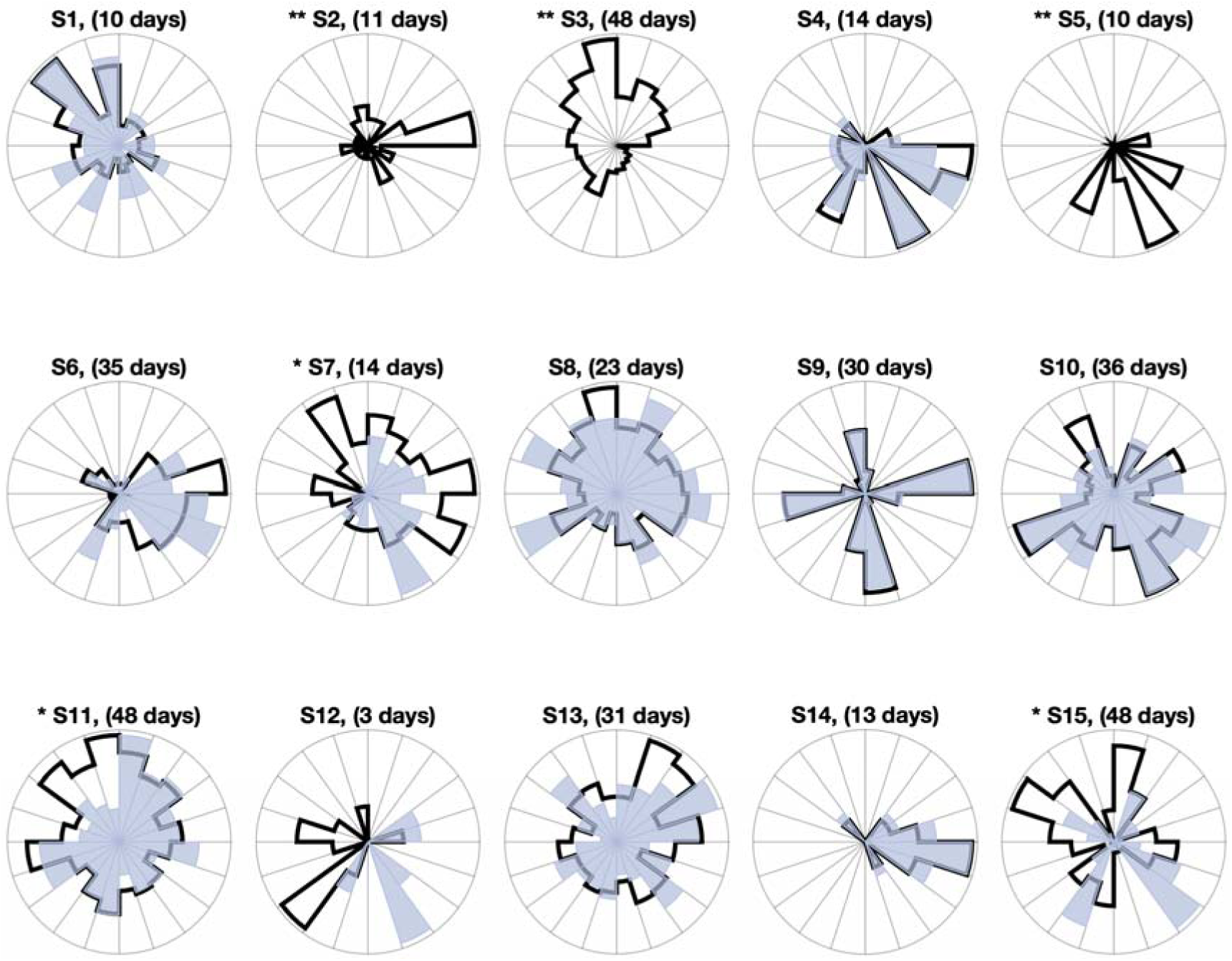
Alignment of seizure cycles based on self-report and EEG. Each circular histogram shows the multiday distribution of seizure cycles based on self-reported events (light blue) and electrographic seizures confirmed by expert reviewers (black line). The period of the multiday cycle for each subject is given above the circular histogram subplots. Subjects 7, 11 and 15 had significantly different cycle distributions using Kuiper’s test for circular distributions (* p < 0.05, ** p < 0.01). Subjects 2, 3 and 5 did not have enough self-reported events for a comparison to be made.

**Figure S2.**
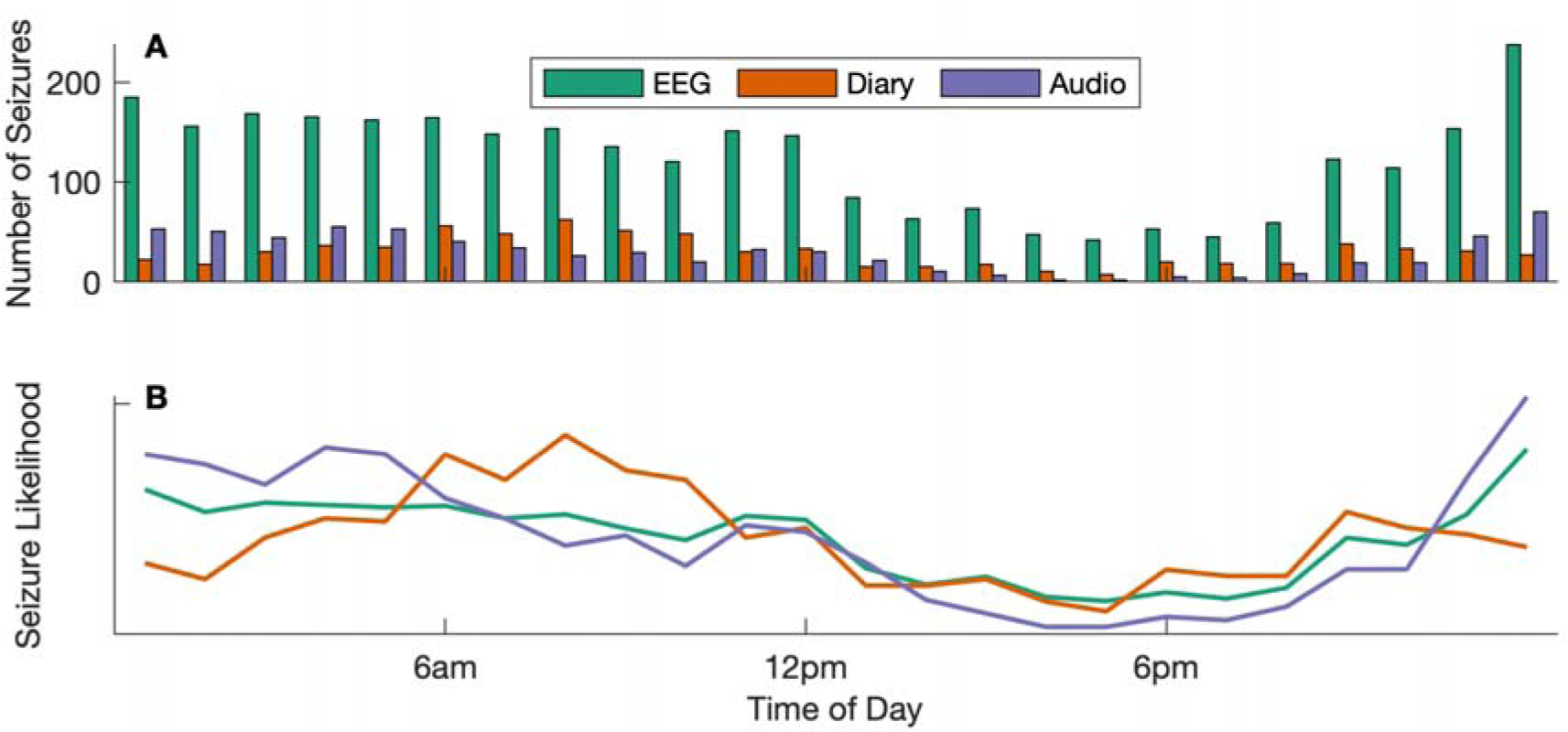
Total seizure rates with respect to time of day. Comparison of total seizure rates and circadian trends across all 15 subjects in the Neurovista cohort. Electrographic seizure rates (“EEG”) are compared to self-reported seizures (“Diary”) and seizures that were confirmed by audio recordings (“Audio”). **A**. Total count of seizures (y-axis) at different hours of day (x-axis). **B**. Circadian trends showing seizure likelihood (y-axis) with respect to time of day (x-axis).

**Figure S3.**
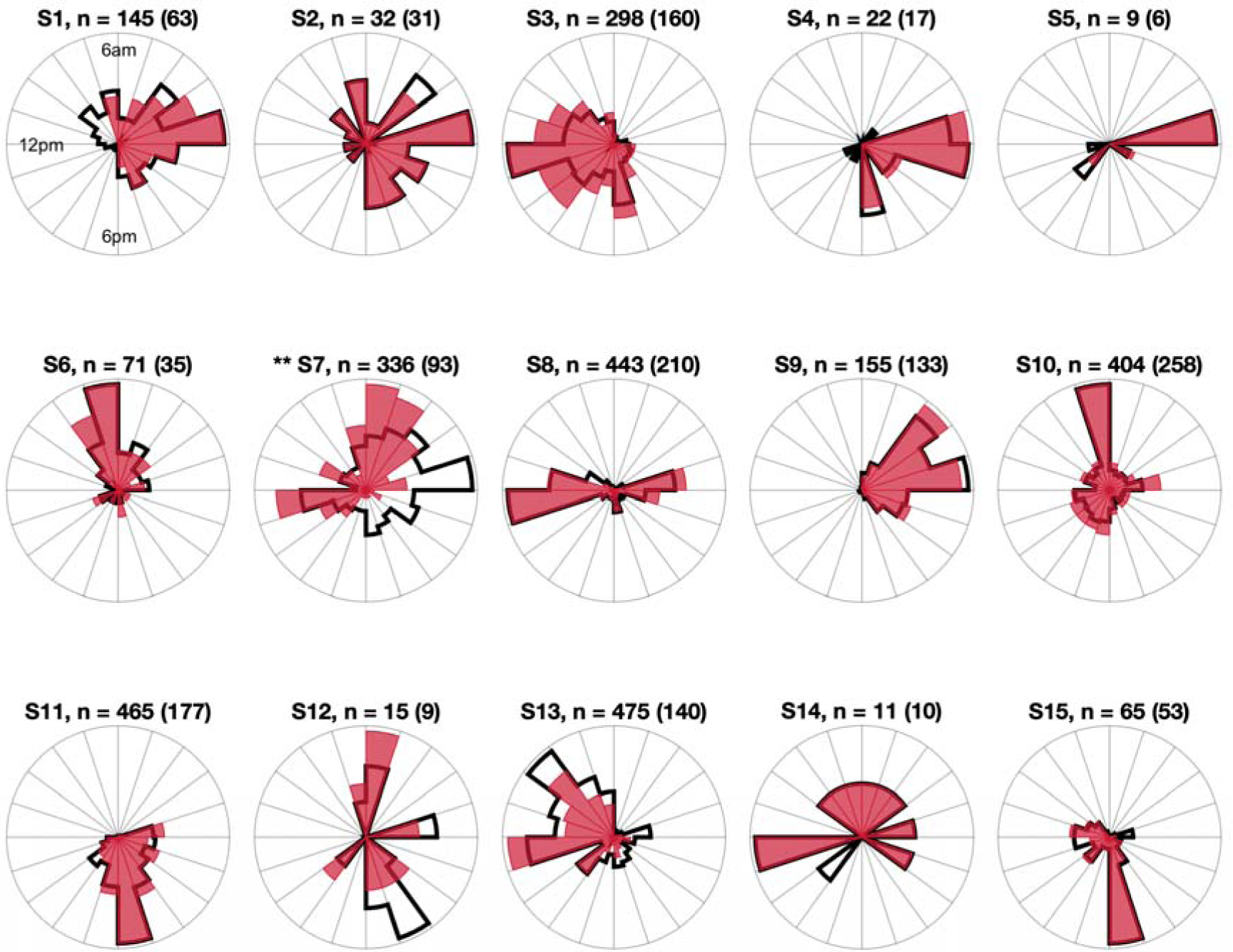
Alignment of seizure cycles based on self-reported + audio detections and EEG. Each circular histogram shows the circadian distribution of seizure cycles based on self-reported events combined with audio confirmed events (pink) and electrographic seizures confirmed by expert reviewers (black line). Total seizure numbers for each subject are given above each plot (with self-reported + audio events in brackets). Subject 7 had significantly different cycle distributions using Kuiper’s test for circular distributions (** p < 0.01).

**Figure S4.**
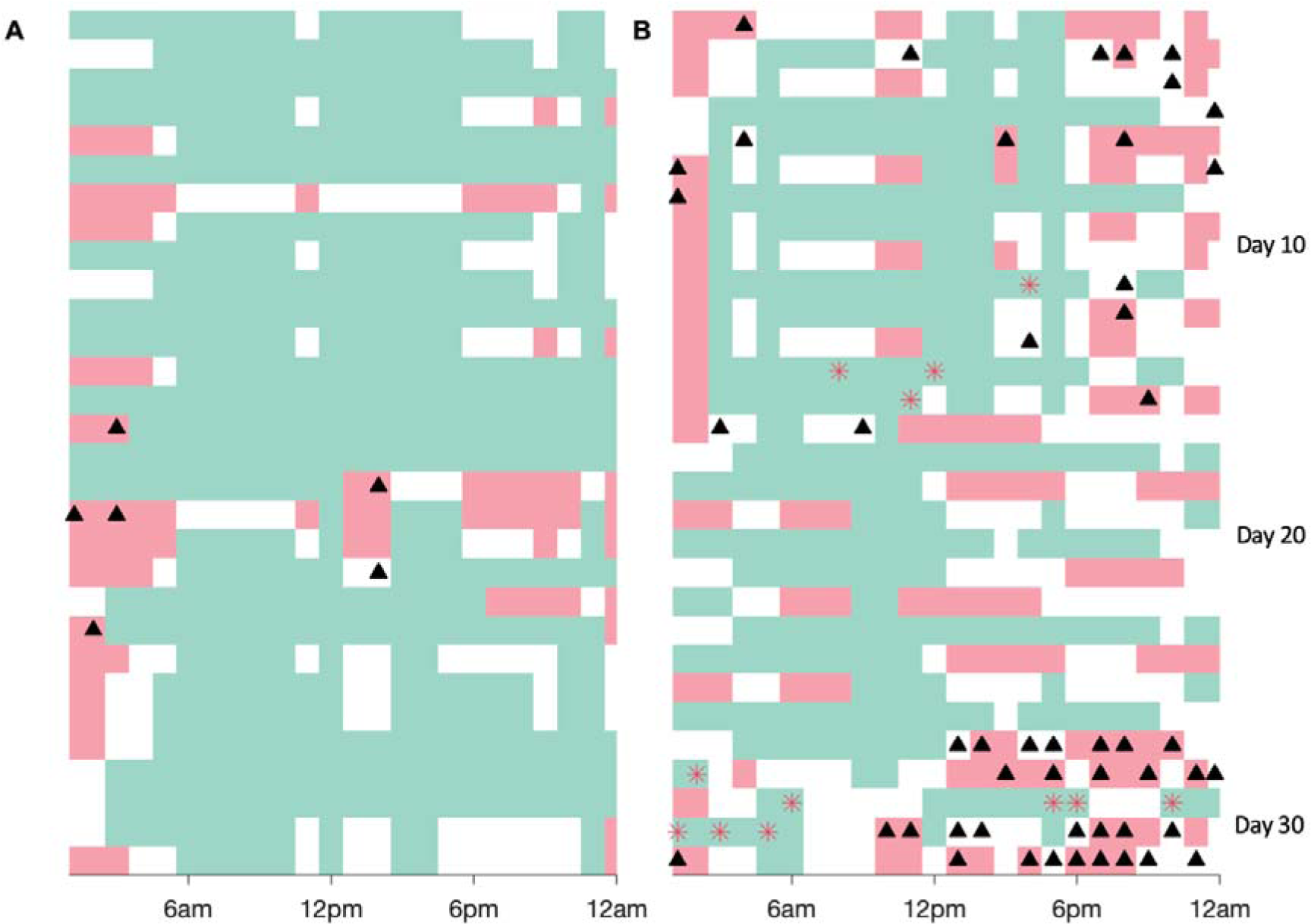
Examples of seizure forecast output. Example output of pseudoprospective seizure forecasts for two different individuals. High risk periods are shown in red, low risk periods are shown in green and moderate risk periods are shown in white. Black triangles represent reported lead seizures (first seizure in a given hour). Red asterisks represent reported seizures that occurred during low risk periods. **A**. A person with a lower reported rate of 1.4 seizures per week; they had no seizures occur during low risk times (53.5% of the time spent in low risk) and 84.6% of seizures during high risk times (23% of the time). **B**. A person with a higher reported rate of 24.2 seizures per week; they had 33 seizures (16.8%) during low risk times (51.7% of the time spent in low risk) and 47.5% of seizures during high risk times (21.9% of the time).

